# Effectiveness of community non-pharmacological interventions for mild cognitive impairment and dementia: a systematic review of economic evaluations and a review of reviews

**DOI:** 10.1101/2022.12.16.22283561

**Authors:** Gillian Eaglestone, Evdoxia Gkaintatzi, Charlotte Stoner, Rosana Pacella, Paul McCrone

## Abstract

**Background:** Dementia prevalence is increasing with no cure at present. Drug therapies have limited efficacy and potential side effects. People with dementia are often offered non-pharmacological interventions to improve quality of life and relieve symptoms. Identifying which interventions are cost-effective is important due to finite resources in healthcare services.

**Aims:** To review published economic evaluations of community non-pharmacological interventions for people with mild cognitive impairment or dementia and assess usefulness for decision making in health services.

**Methods:** Systematic review (PROSPERO CRD42021252999) included economic evaluations of non-pharmacological interventions for dementia or mild cognitive impairment with a narrative approach to data synthesis. Exclusions: interventions for dementia prevention/early detection or end of life care. Databases searched: Academic search premier, MEDLINE, Web of Science, EMBASE, Google Scholar, CINAHL, PsycInfo, Psychology and behavioural sciences collection, PsycArticles, Cochrane Database of Systematic Reviews, Business Source Premier and Regional Business News; timeframe 01 January 2011 to 30 June 2021 (13 September 2021 for Embase). Study quality assessed using CHEERS.

**Results:** Included thirty-two studies and five reviews, evaluating community dementia interventions worldwide across several distinct forms of care: physical activity, cognition, training, multi-disciplinary interventions and other (telecare/assistive technology, specialist dementia care, group living, home care versus care home). No single intervention was shown to be cost-effective across all economic evaluations.

**Conclusion:** More economic evidence on the cost-effectiveness of specific dementia care interventions is needed, with consistency around measurement of costs and outcomes data. Better information and higher-quality studies could improve decision makers’ confidence to promote future cost-effective dementia interventions.

## Introduction

The worldwide economic burden of dementia care is high at US$815 billion (1). The total annual cost of dementia in the UK is estimated at £24.2 billion (2). With the increase in numbers of patients being diagnosed with dementia and the high costs of dementia care, economic evaluations are needed to ensure that non-pharmacological therapies which are offered to patients are cost-effective. However, economic evidence of non-pharmacological dementia interventions remains limited (3).

By 2050 the number of people with dementia is projected to rise to 152 million due to population growth and an increasingly ageing population (4). There is currently no cure for dementia; drug therapies developed to date have limited efficacy and are primarily indicated for use in Alzheimer’s Disease, which may account for 60-70% of cases (5). Probability of cost-effectiveness of drug therapies including rivastigmine and galantamine is low (6) with potential for serious side-effects including mortality (7). Therefore non-pharmacological therapies may be considered as complements to pharmacological treatments.

The National Institute for Health and Care Excellence guideline for dementia care (8) recommends four non-pharmacological interventions: group cognitive stimulation therapy, group reminiscence therapy and cognitive rehabilitation or occupational therapy. The main aim of these types of dementia interventions is to reduce symptoms including cognitive decline, promote independence and wellbeing and improve quality of life.

Existing systematic reviews of economic evidence commonly focus on a particular intervention (9,10) or dementia symptom (11,12). Previous reviews have also included interventions to improve the quality of life of carers as well as people with dementia (PwD) (3,13). The aim of this review was to provide a comprehensive summary of the existing economic evidence on non-pharmacological interventions, evaluating a wide range of dementia symptoms and interventions which measured the impact on the PwD and not solely their carer.

## Methods

The protocol for this systematic review was registered on PROSPERO (CRD42021252999). PRISMA guidelines were followed throughout (14).

### Eligibility Criteria

Inclusion criteria (detailed in Table 1) stated that studies should be economic evaluations, observational studies or simulation studies; the population under observation was people with dementia or mild cognitive impairment. People with mild cognitive impairment were included as a high percentage go on to later develop dementia (15). To be eligible interventions needed to aim to delay progression of the disease or improve quality of life. Studies could have evaluated dementia interventions throughout the dementia pathway, ranging in severity from recent diagnosis to advanced dementia, but prevention/early detection of dementia studies or end of life care studies were excluded. Both narrative and systematic reviews of economic studies were also eligible for inclusion.

**Table 1.** Eligibility Criteria.

| **Inclusion criteria** | **Exclusion criteria** |
| --- | --- |
| Population with dementia (including Alzheimer's disease) or mild cognitive impairment | Studies of intervention purely for the carers of people with dementia |
| Trial based economic evaluations, observational or simulation studies | Interventions focusing solely on homeopathic or herbal remedies |
| Interventions designed to either delay progression of the disease or to improve/maintain health-related quality of life | End of life care interventions |
| All study designs including reviews and systematic reviews | Interventions aimed at prevention/early detection of dementia |
| Papers in any language, both abstracts and full texts will be translated | Studies without any costings data |
|  | Letters, commentaries, study protocol papers and conference abstracts |

### Search Strategy

The databases searched were Academic search premier, Google Scholar, Web of Science, Cochrane Database of Systematic Reviews, MEDLINE, CINAHL, PsycINFO, Psychology and behavioural sciences collection, PsycARTICLES, Business Source Premier and Regional Business News. Studies published between 01^st^ January 2011 and 30^th^ June 2021 (13^th^ September 2021 for EMBASE) were included to search only most recent articles. Articles published in any languages were eligible.

Search terms used within the databases were the following disease specific terms: “dementia”, “Alzheimer’s” and “mild cognitive impairment” combined with the economic terms: “cost*” or “econ*” with additional search terms to identify interventions: “intervention” or “therapy”. The reference lists of primary studies and review articles which met the inclusion criteria were manually searched for other relevant studies for inclusion.

### Study Selection

Titles and abstracts were screened according to inclusion/exclusion criteria by author GE and results were verified by EG. Any disputes were resolved by PM. Full texts of selected articles were retrieved and reviewed by GE and EG.

### Data Extraction

Extracted data included intervention description, participant numbers, follow-up period, study design, economic evaluation type, main economic outcome measure, primary outcome (PwD only) and perspective (see Table 2). Data extraction was performed by GE, with EG independently undertaking data extraction for 40% of the included articles. Any disagreement was resolved by PM.

**Table 2.** Study Characteristics.

| **Author, Country** | **Int** | **Intervention Description** | **N** | **F/up mont hs** | **Design** | **EE** | **Main economic outcome measure** | **Primary Outcome** | **Perspective** |
| --- | --- | --- | --- | --- | --- | --- | --- | --- | --- |
| Brown et al (18), UK | Cog | Maintenance cognitive stimulation therapy with a group that had previously received cognitive stimulation therapy. | INT 123, TAU 113 | 6 | RCT | CUA | QALYS | EQ-5Q (self and proxy), DEMQOL (self and proxy) | Societal |
| D'Amico et al (22), UK | Cog | Maintenance cognitive stimulation therapy in addition to usual care. | INT 64 dyads, TAU 67 dyads | 6 | RCT | CUA | QALYS | ADAS-Cog, QoL-AD | H&SC |
| Laakkonen et al (34), Finland | Cog | Separate group rehabilitation sessions run for people with dementia and for spouses. | INT 67 TAU 69 | 24 | RCT | CEA | Patient health related quality of life | 15D | Not stated |
| Mervin et al (44), Australia | Cog | Individual, non-facilitated sessions with a soft toy. Groups received toy with artificial intelligence (PARO) or same toy (Plush toy) with artificial intelligence disabled. | Paro 138, Plush 140, TAU 137 | 2.5 | RCT | CEA | Patient agitation level | CMAI-SF | HCP |
| Orgeta et al (23), UK | Cog | Home-based, individual cognitive stimulation therapy, led by own carer. Up to three 30-minute activity sessions/week over 25 weeks. Initial training and ongoing support provided. | INT 180, TAU 176 | 25 | RCT | CUA | QALYS | ADAS-Cog, QoL-AD | H&SC/ Societal |
| Sado et al | Cog | A learning therapy intervention | INT 30, | 12 | Non- | CBA | Criterion Time | CT for CNLTC | HCP |
| (45), Japan |  | carried out in nursing homes. A combination of cognitive training and stimulation. | TAU 27 |  | randomised matched control |  | for Certification of Needed Long-Term Care |  |  |
| Sogaard et al (42), Denmark | Cog | Intensive, multi-component, semi-tailored, psychosocial intervention programme with counselling, education and support. | INT 163 dyads<br>TAU 167 dyads | 36 | RCT | CUA | QALYS | QALYS (measured by EQ-5D-5L) | Societal |
| Spector et al (24), UK | Cog | A cognitive behavioural therapy (CBT) manual developed for anxiety in dementia and trialled in sessions with participant/carer dyads. | INT 25,<br>TAU 25 | 6 | RCT | CEA | Patient anxiety | RAID score | H&SC |
| Woods et al (25), UK | Cog | Group reminiscence therapy for participants and with carers. | INT 206 dyads<br>TAU 144 dyads | 10 | RCT | CUA | QALYS | QoL-AD, GHQ-28 (carer) | Public sector |
| Ballard et al (26), UK | Train | Staff training programme on person-centred care for care staff, promoting tailored activities, antipsychotic medication review. | INT 404,<br>TAU 443 | 9 | RCT | CEA | Patient quality of life | DEMQOL- (Proxy) | Not stated |
| El Alili et al (37), Netherlands | Train | Multi-dimensional care programme for nursing home residents with advanced dementia. | INT 116,<br>TAU 115 | 12 | RCT | CUA | QALYS | QUALID, GAIN | Societal |
| Livingston et al (27), UK | Train | Evidence-based, manualised, intervention, delivered to staff in 6 interactive sessions. | INT 189,<br>TAU 215 | 8 | Cluster RCT | CUA | QALYS | CMAI | Not stated |
|  |  | Comprising training getting to know and understand residents. |  |  |  |  |  |  |  |
| Meads et al (7), UK | Train | Dementia Care Mapping: a care-home staff training intervention aimed to embed person-centred care, improve care quality and health outcomes for residents. | INT 418, TAU 308 | 16 | RCT | CUA | QALYS | QALYS (measured by EQ-5D-5L) | HCP |
| Menn et al (30), Germany | Train | GPs in 3 groups (A, B, C) trained in dementia care. At different timepoints groups B and C actively recommended support groups and promoted caregiver counselling. | B 88, C 108, TAU 107 | 48 | RCT | CEA | Time to institutionalisation | Remaining time the patient lived at home | Societal |
| Romeo et al (21), UK | Train | Person-centred care intervention delivered by staff in nursing homes. Staff were trained in person-centred care, management of agitation, and psychosocial approaches. | INT 404, TAU 443 | 9 | RCT | CUA | QALYS | QOL measured by DEMQOL | H&SC |
| Van de Ven et al (38), Netherlands | Train | Dementia-care mapping, multi-component intervention. Staff from participating nursing homes attended training, then assessed residents' needs and implemented care interventions. | INT 154, TAU 164 | 18 | RCT | CBA | Health care consumption, falls and psychotropic drug use and staff absenteeism | Agitation | HCS |
| Williams et al (40), USA | Train | Staff training intervention to improve communication with residents by encouraging staff to stop using 'childish' | INT 42 dyads, TAU unstated | 36 | Observational | CEA | % Time staff used Elderspeak, % time residents | % Time staff used Elderspeak, % time residents | not stated |
|  |  | language/way of speaking and interacting (Elderspeak). |  |  |  |  | resistive to care | resistive to care |  |
| Henderson et al (48), Italy, Poland, UK | Multi-disciplinary | Meeting centre support programme; person-centred, psychosocial approach combining day centre services with support for carers. | INT 83, TAU 69 | 6 | Non-randomised matched control | CUA | QALYS | QOL-AD | H&SC /Societal |
| Jennings et al (41), USA | Multi-disciplinary | Dementia care programme co-managed by nurse practitioners and physicians, comprising structured assessments, individualised care plans, referral to community support/advice services, and 24hr access to a clinician. | INT 1083, TAU 2186 | 36 | RCT | CEA | Patient health care utilisation | Admission to long-term care, healthcare utilisation | Not stated |
| MacNeil Vroomen et al (39), Netherlands | Multi-disciplinary | Comparison of 2 types of case management. Intensive Case Management Model: carried out within one care organization, and Linkage Model where care was provided by different care organisations within 1 region. | Intensive 234, Linkage 214, TAU 73 | 24 | Non-randomised observational controlled, cohort study | CUA | QALYS | NPI score | Societal |
| Michalowsky et al (32), Germany | Multi-disciplinary | Dementia care management intervention using multi-disciplinary care to assess and support the dementia patient and carer. | INT 315, TAU 129 | 24 | RCT | CUA | QALYS | SF-12 | Societal |
| Mostardt et al (31), Germany | Multi-disciplinary | Case management intervention aiming to improve support networks and quality of care and enable PwD to stay at home for longer. | INT 38, TAU 76 | 12 | Non-randomised matched control | CEA | Additional time in the home environment | Time in the home environment | HCP |
| Rädke et al (33), Germany | Multi-disciplinary | Dementia care management intervention, delivered in participant's homes by dementia-trained nurses. Focus on interprofessional treatment and care, medication management and support/education of carers. | INT 315, TAU 129 | 24 | Cluster randomised | CEA | Patient functional independence | SF-12 | HCS |
| Rosenvall et al (35), Finland | Multi-disciplinary | Three interventions for people with Alzheimer's: care-management (CM), family support (FS) and physical activity (P). Analysed using existing data then a modelling simulation was carried out. | 924 (simulation) | 48 | Modelling | CBA | Use of health and care services, clinical outcome measures | Delay in transition to long term care | Not stated |
| Saxena et al (47), Singapore | Multi-disciplinary | Primary care dementia clinic (PCDC) compared with a hospital-based memory clinic (MC). PCDC was run by GPs and nurses assisted by the MC team. | MC 101, PCDC 99, TAU 63 | 12 | Non-randomised quasi-experimental design | CUA | QALYS | QALYS (measured by EQ-5D-5L) | Societal |
| D'Amico et al (20), UK | PE | Individually tailored dyadic regime, walking daily for 12 weeks. | INT 30, TAU 22 | 3 | RCT | CUA | QALYS | NPI score | Societal |
| Davis et al (46), Canada | PE | Twice-weekly group classes for older women, comparing resistance or aerobic training with balance and tone classes. | Resistance 28, Aerobic 30, Balance & Tone 28 | 6 | RCT | CEA | Patient executive cognitive function | Stroop test | HCP |
| Lamb et al (28), UK | PE | Weekly group exercise programme of aerobic and resistance training led by a physio. | INT 329, TAU 165 | 12 | RCT | CUA | QALYS | ADAS-Cog score | HCP/ Societal |
| Pitkala et al (36), Finland | PE | Group-based exercise sessions twice a week compared to home-based exercise. | Home 70, Group 70, TAU 70 | 12 | RCT | CEA | FIM score changes and SPPB score changes | FIM, SPPB | H&SC |
| Sopina et al (43), Denmark | PE | Supervised, hour-long group sessions of aerobic exercise, three times a week. | INT 107, TAU 93 | 4 | RCT | CUA | QALYS | Symbol Digit Modalities Test | Danish healthcare perspective |
| Howard et al (29), UK | Other | Assistive technology and telecare assessment followed by installation of all appropriate devices. | INT 248, TAU 247 | 24 | RCT | CUA | QALYS | Time to institution-alisation | H&SC /Societal |
| Tanajewski et al (19), UK | Other | A specialist unit was developed within a general hospital, for older people admitted for acute medical care assessed as having delirium and dementia. | INT 309, TAU 290 | 3 | RCT | CUA | QALYS | QALYS (measured by EQ-5D-5L) | H&SC |

**Key:** ADAS-Cog - Alzheimer's Disease Assessment Scale–Cognitive Subscale, Cat – Category, CBA – cost-benefit analysis, CEA – cost-effectiveness analysis, CUA – cost-utility analysis, DEMQOL - Dementia Quality of Life, EE – method of economic evaluation, EQ-5D - health related quality of life measure, EQ-VAS - EQ-Visual Analogue Scale, F/up – length of follow up in months, FIM - Functional Independence Measure, GAIN - Alzheimer's care instrument, H&SC – health and social care, HCP – healthcare provider, HRQoL - health-related quality of life, HS – health service, INT - intervention, N – number in sample, NPI - neuropsychiatric Inventory, PE – physical exercise, QALYS – quality adjusted life years, QOL – quality of life, QoL-AD - Quality of Life in Alzheimer Disease, QUALID - Quality of life in late stage dementia, RCT – randomised controlled trial, SF-12 - Short Form Health Survey, SPPB - Short Physical Performance Battery, TAU – treatment as usual

Separate data extraction was undertaken for the review of reviews by GE (see Table 3). Extracted data included: interventions reviewed, number of studies included in the review and databases used.

**Table 3.** Review Characteristics.

| **Study** | **Review Method** | **Interventions reviewed** | **Intervention Descriptions** | **No. studies included** | **Databases Used** |
| --- | --- | --- | --- | --- | --- |
| Alves et al (9) | Systematic | Cognitive | Cognitive stimulation, cognitive training or cognitive rehabilitation | 5 | PubMed, PsychINFO, The Cochrane Library, EMBASE, metaRegister of Clinical Trials, OVID, EBM Reviews |
| Banerjee (50) | Narrative | Multi-disciplinary | Memory services | Not stated | Not stated |
| Clarkson et al (13) | Systematic | Multi-disciplinary | Home support interventions | 8 | British National Health Service Economic Evaluation Database (NHSEED) |
| Livingston et al (11) | Systematic | Cognitive/Multi-disciplinary | Blended inpatient and outpatient programme to reduce hospitalisation. Also an Australian comparison of dementia care mapping and person-centred care programmes. | 2 | MEDLINE, Web of Knowledge; EMBASE; British Nursing Index; the Health Technology, NHS Economic Evaluation Database, the HTA programme database and the DARE Assessment programme database; PsycINFO; NHS Evidence; System for Information on Grey Literature; The Stationery Office Official Documents website; The Stationery National Technical Information Service; Cumulative Index to Nursing and Allied Health Literature; and The Cochrane Library. |
| Nickel et al (3) | Systematic | Physical exercise/Cognitive | Physical exercise, cognitive interventions | 16 | Centre for Reviews and Dissemination, EconLit, Embase, Cochrane Library, PsycINFO and PubMed |

### Data Synthesis

A narrative approach to data synthesis was undertaken to summarise and allow comparison of the methods and results of the included studies whilst demonstrating heterogeneity. A narrative reporting approach was used as meta-analysis could not be carried out due to the context specific nature of the economic evaluations and the numerous outcomes used in these studies.

Interventions were classified according to the following categories representing distinct forms of care: physical activity, cognitive interventions, training interventions, multi-disciplinary interventions, other interventions (including telecare/assistive technology, specialist dementia care, group living, home care versus care home).

### Quality Appraisal

Quality appraisal was undertaken using the Consolidated Health Economic Evaluation Reporting Standards statement (CHEERS) (16). CHEERS is designed to assess reporting quality rather than the quality of the study. The statements relate to the following aspects: title, abstract, introduction, methods, results, discussion and disclosure. Statements which related only to modelling studies were excluded for non-modelling studies and resulting scores adjusted accordingly. Each study was assigned a score, based on the number of statements met on the CHEERS checklist (0 = unmet, 0.5 = partially met, and 1 = met), the total was then translated into a percentage of items met. Quality appraisal was carried out by GE, EG undertook an independent appraisal of 30% of the articles and any disagreement was resolved by PM.

For the review of reviews, quality was assessed by GE using A Measurement Tool to Assess Systematic Reviews (AMSTAR 2) (17) with EG independently undertaking assessment for 60% of the included articles (see Table 4). The AMSTAR 2 assessment tool is intended to identify areas of potential bias by assessing each area of the study, with certain areas being defined as critical. An overall rating of confidence in quality was calculated based on the total number of unmet statements on the AMSTAR 2 checklist according to the following criteria:

- *High confidence* - none or 1 non-critical weaknesses,
- *Moderate confidence -* more than 1 non-critical weakness but no critical flaws,
- *Low confidence* - 1 critical flaw with or without non-critical weaknesses,
- *Critically low confidence* - more than 1 critical flaw with or without non-critical weaknesses.

**Table 4.**
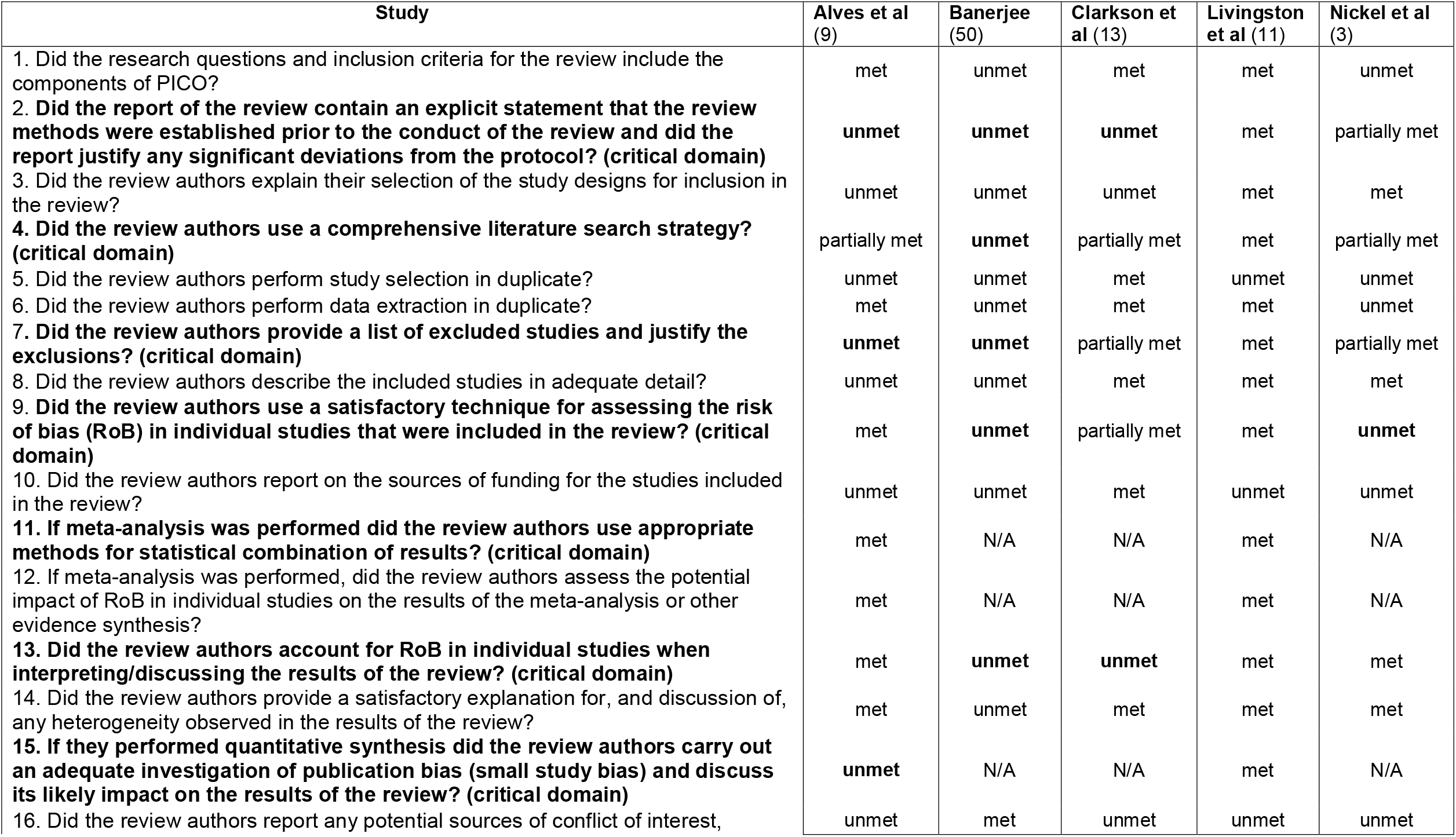
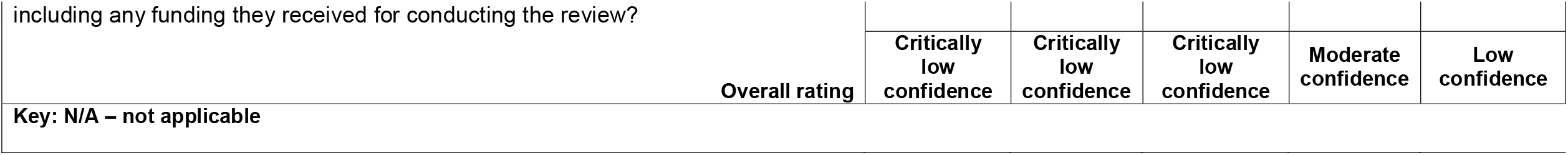
Quality Assessment of Systematic Reviews using AMSTAR 2 (17)

### Usefulness of Economic Evaluations to Decision Making

A score of usefulness of the economic evaluations to decision making was calculated for each included study. It was based on assessment of reporting quality, study design, time horizon (above or below 12 months) and whether an ICER was reported. Studies were then categorised as having a ‘strong’, ‘moderate’ or ‘limited’ level of usefulness (see Table 5).

**Table 5.** Usefulness of Economic Evaluations to Decision Making.

| **Study** | **Level of usefulness** | **Primary reasons** |
| --- | --- | --- |
| MacNeil Vroomen et al (39) | Strong | Appropriate statistical analysis, time horizon > than one year |
| Meads et al (7) | Strong | Appropriate statistical analysis, time horizon > than one year |
| Michalowsky et al (32) | Strong | Appropriate statistical analysis, time horizon > than one year |
| Orgeta et al (23) | Strong | Appropriate statistical analysis, time horizon > than one year |
| Rädke et al (33) | Strong | Appropriate statistical analysis, time horizon > than one year |
| Tanajewski et al (19) | Strong | Appropriate statistical analysis, time horizon > than one year |
| Rosenvall et al (35) | Strong | Sensitivity analysis performed, 4-year time horizon |
| El Alili et al (37) | Strong | Appropriate statistical analysis, time horizon one year |
| Lamb et al (28) | Strong | Appropriate statistical analysis, time horizon one year |
| Brown et al (18) | Moderate | Short time horizon, ICER was reported |
| Laakkonen et al (34) | Moderate | Long time horizon, ICER not reported |
| D'Amico et al (22) | Moderate | Appropriate statistical analysis, short time horizon |
| D'Amico et al (20) | Moderate | Appropriate statistical analysis, short time horizon |
| Howard et al (29) | Moderate | Appropriate statistical analysis, short time horizon |
| Livingston et al (27) | Moderate | Appropriate statistical analysis, short time horizon |
| Pitkala et al (36) | Moderate | Appropriate statistical analysis, no ICER reported |
| Romeo et al (21) | Moderate | Appropriate statistical analysis, short time horizon |
| Sogaard et al (42) | Moderate | Appropriate statistical analysis, no ICER presented |
| Sopina et al (43) | Moderate | Appropriate statistical analysis, short time horizon |
| Woods et al (25) | Moderate | Appropriate statistical analysis, short time horizon |
| Mostardt et al (31) | Moderate | Appropriate statistical analysis, not an RCT or observational study |
| Williams et al (40) | Moderate | Statistical analysis not appropriate, time horizon > than one year |
| Saxena et al (47) | Limited | ICER not reported for all comparison groups |
| Menn et al (30) | Limited | Statistical analysis not appropriate, no ICER presented |
| Ballard et al (26) | Limited | Short time horizon, no ICER presented |
| Davis et al (46) | Limited | Short time horizon, no ICER presented |
| Jennings et al (41) | Limited | Analysis of uncertainty not stated, no ICER presented |
| Spector et al (24) | Limited | Short time horizon, no ICER presented |
| Van de Ven et al (38) | Limited | Statistical analysis not appropriate, no ICER presented |
| Henderson et al (48) | Limited | Short time horizon, not an RCT or observational study |
| Sado et al (45) | Limited | No ICER presented, not an RCT or observational study |
| Mervin et al (44) | Limited | Short time horizon, no ICER presented, lack of sensitivity analysis |

## Results

The systematic literature review identified 664 publications, duplicates were manually removed, and 405 articles were screened. Thirty-seven papers were included in the final review. The articles selected for inclusion comprised 32 single study papers and five reviews. The search strategy has been reported using the Preferred reporting items for systematic review and meta-analysis (PRISMA) (14) flow diagram (Figure 1).

**Figure 1.**
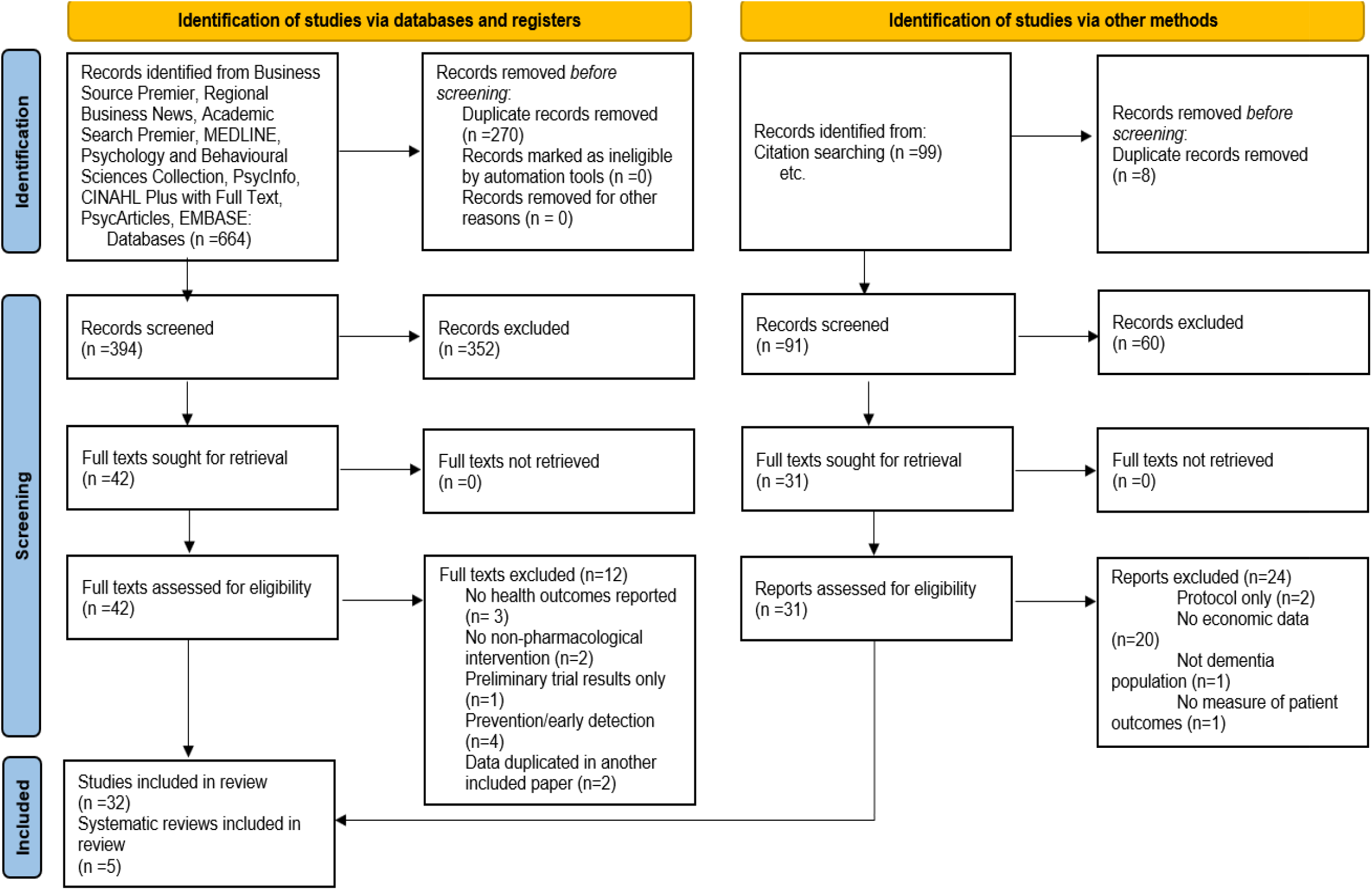
PRISMA flow diagram (14)

### Study Characteristics

The specific conditions being studied included: people with dementia (n=26), Alzheimer’s Disease (n=2), MCI (n=1), and mixed populations which included MCI, delirium and dementia (n=3). Types of interventions reviewed were: exercise (n=5), cognitive (n=9), multi-disciplinary (n=8), training (n=8) and other: telehealth/assistive technology (n=1), specialist dementia care unit (n=1). Studies recruited participants living in a variety of settings: nine were community based, nine were nursing home based, three were for people living either in a nursing home or the community and one was unspecified. Studies used varying criteria to define dementia/MCI in their inclusion criteria, ranging from a having symptoms of dementia (n=1) to a formal diagnosis of dementia (n=8). A number of studies defined specific Clinical Dementia Scores (n=3) and/or Diagnostic and Statistical Manual of Mental Disorders (DSM-IV) assessment criteria in their inclusion criteria (n=6).

The majority of studies were randomised controlled trials (n=26), followed by non-randomised studies (n=5) and one modelling study. Study sample size varied greatly, ranging from 50 to 3269. International reach of the studies was the UK (n=13) (7,18–29), Germany (n=4) (30–33), Finland (n=3) (34–36) and Netherlands (n=3) (37–39), United States (n=2) (40,41)and Denmark (n=2) (42,43), Australia (n=1) (44), Japan (n=1) (45), Canada (n=1) (46), Singapore (n=1 (47) and there was a multi-national study involving Italy, Poland and the UK (48).

A range of interventions were identified, which focused on improving the behavioural and psychological symptoms of dementia (BPSD), as well as interventions to prevent decline in cognitive function and mobility. In total 27 different primary outcome measures were used across the 33 studies. Outcomes included generic, dementia specific and utility-based quality of life scales.

Over half of the studies employed a cost utility analysis (n=18), followed by cost-effectiveness analysis (n=11) and cost benefit analysis (n=3), QALY was the most frequently used measure of benefit (n=18). Costs and outcomes are reported in Table 6.

**Table 6.** Study Results.

| Author, Country | Int | Intervention group cost<sup>a</sup> | Comparison group cost | Intervention group outcome | Comparison group outcome | Main result |
| --- | --- | --- | --- | --- | --- | --- |
| Brown et al (18), UK | Cog | Care Home £18,635; Lives alone £9,053; lives with someone £18,853; Low ADAS-Cog 18,675; High ADAS-Cog £15,266 | Care Home £17,485; Lives alone £9,155; lives with someone £15,026; Low ADAS-Cog £16,398; High ADAS-Cog £13,791 | 0.07 QALYS (EQ-5D) Lives alone |  | Intervention dominant over TAU in terms of QALYS for the sub-group living alone |
| D'Amico et al (22), UK | Cog | £11,306 | £11,440 |  | Incremental effect: 0.0176 QALYS (calculated with Proxy EQ-5D) | ICER = £26,835/QALYS (calculated by Proxy EQ-5D) |
| Laakkonen et al (34), Finland | Cog | €8,947 | €9,383 | -0.03 HRQoL mean score | -0.04 mean change in HRQoL | There was no change between the groups in HRQoL according to 15D. However, the intervention had beneficial effects on cognitive function without increasing total costs |
| Mervin et al (44), Australia | Cog | \$13,827 (PARO, interactive toy); \$12,078 (Soft toy) | \$6,862 | -2.66 CMAI-SF mean change (PARO), -1.68 CMAI-SF mean change (Soft toy) | 1.22 CMAI-SF mean change | Incremental cost per unit improvement in CMAI-SF was \$13.01 for the PARO group and \$12.85 for the soft toy group relative to usual care |
| Orgeta et al (23), UK | Cog | £4,740 (HSC); £9770 (Societal) | £4,670 (HSC); £10,630 (Societal) | ADAS-Cog 20.53, QoL-AD 37.96 | 21.19 ADAS-Cog, 37.71 QoL-AD | Intervention dominated by TAU in terms of ADAS-Cog, no differences in primary outcomes of cognition or quality of life |
| Sado et al (45), Japan | Cog | \$795 | not stated | 68.4 CT for CNLTC mean | 83.3 mean score CT for | Intervention showed \$21 per month cost-saving/unit of CT for CNLTC for avoiding |
|  |  |  |  | score | CNLTC | increased long term care costs |
| Sogaard et al (42), Denmark | Cog | € 84,142 per dyad | € 80,741 | 3.26 QALYS | 3.46 QALYS | Intervention dominated by TAU in terms of QALYS |
| Spector et al (24), UK | Cog | £2,344 | £1,259 | -4.59 mean score<br>Rating anxiety in dementia for PwD |  | Intervention was cost neutral |
| Woods et al (25), UK | Cog | £5,853 | £4,309 | 37.013 QoL-AD, 0.644 QALYS | 36.416 QoL-AD, 0.643 QALYS | ICER = £2,586/per 0.597 change in QoL-AD score (no ICER expressed in £/QALYS) |
| Ballard et al (26), UK | Train | £2,713 (intervention cost); £29702 (total HSC costs) | £0 (intervention cost); £34,442 (total HSC costs) | 4.78-point improvement in DEMQOL | 2.54-point improvement in DEMQOL (mean difference) | Intervention demonstrates benefits in terms of quality of life, agitation, and neuropsychiatric symptoms, as well as cost savings |
| El Alili et al (37), Netherlands | Train | £7,173 (total societal costs) | £7,484 (total societal costs) | Incremental effect: 0.40 QALYS | 0.39 QALYS | Intervention dominant over TAU in terms of QALYS |
| Livingston et al (27), UK | Train | £1,379 | £1,175 | 0.346 QALYS | 0.332 QALYS | ICER = £14,064/QALYS gained |
| Meads et al (7), UK | Train | £3,539 | £2,060 | 0.718 QALYS | 0.708 QALYS | ICER = £64,380/QALYS gained |
| Menn et al (30), Germany | Train | €80,361 (Group B); €75,754 (Group C) | Group A €82,745 | x | x | Group B = 0.858 Hazard rate on time to institutionalisation over 4 years<br>Group C = 1.133 Hazard rate on time to institutionalisation over 4 years |
| Romeo et al (21), UK | Train | £32,112 | £34,215 | 1.96 DEMQOL proxy score (mean change) |  | Intervention dominant over TAU in terms of QALYS gained |
| Van de Ven et al (38), Netherlands | Train | \$0.63 per resident/day | not stated | 3.33 falls (mean annual no.), \$3.50 per resident/day (total healthcare consumption), \$0.13 per | 1.81 falls (mean annual no.), \$3.23 per resident/day (total healthcare consumption), \$0.25 per | Intervention was cost neutral |

|  |  |  |  | resident/day<br>(psychotropic<br>drug use) | resident/day<br>(psychotropic<br>drug use) |  |
| --- | --- | --- | --- | --- | --- | --- |
| Williams et al (40),<br>USA | Train | \$79.69/per staff<br>member<br>(Elderspeak);<br>\$39.84/per<br>resident<br>(Resistive to<br>care) | x | 11.80%<br>(reduction in time<br>staff used<br>Elderspeak), 9.24<br>% reduction in<br>time residents<br>resistive to care | | ICER = \$4/resident per one percentage point<br>reduction in Resistiveness to care<br>ICER = \$7/staff member per one percentage<br>point reduction in Elderspeak |
| Henderson et al<br>(48), Italy, Poland,<br>UK | Multi-<br>discipl<br>inary | £10,650 (total<br>health & social<br>care costs) | £4,709 (total<br>health & social<br>care costs) | Incremental effect<br>0.40 QALYS | 0.40 QALYS,<br>mean difference<br>0.01 QALYS | ICER = £832,636/QALYS gained for PwD.<br>For QALYS the probability of c/e was 0 at<br>WTP of £0 to £350,000. For QOL-AD, the<br>probability of c/e of intervention was 50% at<br>WTP of €5,000 for a one-point increase |
| Jennings et al (41),<br>USA | Multi-<br>discipl<br>inary | \$317 per quarter | not stated | not stated | not stated | Hazard ratio of 0.60, program participants<br>less likely to be admitted to long-term care<br>facility than TAU |
| MacNeil Vroomen<br>et al (39),<br>Netherlands | Multi-<br>discipl<br>inary | €69,435<br>(ICMM);<br>€84,155 (LM) | €107,627 | 1.25 QALYS<br>(ICMM), 1.18<br>QALYS (LM) | 1.27 QALYS | ICER (ICMM v LM) = ICMM dominates<br>ICER (LM v control) = €460,135<br>ICER (ICMM v control) = unstated |
| Michalowsky et al<br>(32), Germany | Multi-<br>discipl<br>inary | €24,046 | € 24,615 | 1.349 QALYS | 1.300 QALYS | Intervention dominant over TAU in terms of<br>QALYS |
| Mostardt et al (31),<br>Germany | Multi-<br>discipl<br>inary | €1,409 | € 1,292 | 16.4 (months in<br>home<br>environment) | 12.2 (months in<br>home<br>environment) | ICER= €53 per additional month in a home<br>environment |
| Rädke et al (33),<br>Germany | Multi-<br>discipl<br>inary | unavailable | unavailable | +0.05 QALYS | Incremental<br>effect: +0.049<br>QALYS | Intervention dominant over TAU in terms of<br>QALYS gained |
| Rosenvall et al<br>(35), UK | Multi-<br>discipl<br>inary | €527 (CM);<br>€8,000<br>(Rehabilitative);<br>€341 (SFS) | N/A |  | N/A | Economic break-even points reached if a<br>patient's transition to long-term care is<br>delayed by:<br>Care management = 2.8 days |
|  |  | All costs per pt/year |  |  |  | Structured family support = 1.8 days<br>Rehabilitative cognitive, physical and social activation = 43.0 days |
| Saxena et al (47), Singapore | Multi-disciplinary | \$15,308 (MC);<br>\$13,275 (PCDC);<br>\$15,766 (Other clinics) | | 0.78 (PCDC),<br>0.76 (Other polyclinics clinics) (EQ-5D scores) | | ICER = \$29,042/QALYS for the PCDC group when compared with the MC group |
| D'Amico et al (20), UK | PE | £10,533 | £7,805 | N/A | Incremental effect: 0.0055 QALYS | ICER = £286,440 QALYS (calculated by DEMQOL-Proxy) Not c/e when outcomes are calculated by QALYS |
| Davis et al (46), Canada | PE | \$2,255 (CAD) (Resistance);<br>\$1,417.15 (CAD) (Aerobic) | \$2,387 (CAD) (Balance and tone) | 44.61 (Resistance),<br>48.27 (Aerobic) (seconds gained/lost on Stroop test) | 54.69 (Balance and tone) (seconds gained/lost on Stroop test) | Both the aerobic and the resistance classes dominate balance and tone classes in terms of seconds gained/lost on Stroop test |
| Lamb et al (28), UK | PE | £5,580 | £3,917 | 0.787 QALYS | 0.826 QALYS | Intervention dominated by TAU in terms of QALYS |
| Pitkala et al (36), Finland | PE | \$25,112 per dyad/per year (HE group);<br>\$2,2066 per dyad/per year (GE group) | \$34,121per dyad/per year (Control group) | -7.1 (HE group),<br>-10.3 (GE group) (FIM change) | -14.4 (control group) (FIM change) | HE & GE interventions had beneficial effects on the physical functioning of pts with AD, without increasing the total costs of health and social services |
| Sopina et al (43), Denmark | PE | £608 (including transport) | Not stated | Not stated | Not stated | ICER= €158,520/QALYS gained |
| Howard et al (29), UK | Other | £19,649 (HSC),<br>£56,000 (Societal) | £15,186 (HSC),<br>£53,378 (Societal) | 1.201 QALYS | 1.306 QALYS | ICER= £8,635/QALYS (HSC),<br>£33,672/QALYS (Societal) |
| Tanajewski et al (19), UK | Other | £7,714 | £7,862 | 0.109 QALYS | 0.108 QALYS | Intervention dominant over TAU in terms of QALYS |
| <sup>a</sup> - Mean cost per person over the length of the intervention unless otherwise stated. |  |  |  |  |  |  |

**Key:** ADAS-Cog - Alzheimer's Disease Assessment Scale–Cognitive Subscale, CAD – Canadian dollars, C/E – cost effectiveness, CMAI-SF - Cohen-Mansfield Agitation Inventory – short form, CM – Care management, Cog – cognitive, Collab – multi-disciplinary, CT for CNLTC - Criterion Time for Certification of Needed Long-Term-Care, DEMQOL - Dementia Quality of Life, EQ-5D – health related quality of life measure, FIM - Functional Independence Measure, GE group – group-based exercise, HE group – tailored home-based exercise, HRQol - Health related quality of life, HSC – health and social care, ICER – incremental cost effectiveness ratio, ICMM – intensive care management model, INT – intervention type, LM – linkage model, MC – memory clinic, PCDC – primary care dementia clinic, PE – physical exercise, PwD – person with dementia, QALYS – quality adjusted life years, QoL-AD - Quality of Life in Alzheimer Disease, RTC – resistiveness to care, SFS – Structured family support, TAU – treatment as usual, Train – training interventions, WTP – willingness to pay

### Quality Appraisal of Studies

Individual studies met between 62% and 98% of CHEERS quality assessment criteria items (mean 78%). Nineteen studies met over 80% of total assessed items. The studies that evaluated exercise were rated highest for quality with four of the five studies meeting above 80% of items. Evaluations of training interventions scored the lowest overall for number of items met. Although the “other” category met the highest number of items overall at 90% it should be noted that this category evaluated two completely different interventions.

### Usefulness of Economic Evaluations to Decision Making

Scores for level of usefulness were as follows: studies rated as having strong usefulness (n=9), moderate (n=13) and limited (n=10). The results showed that a high CHEERS quality assessment score did not necessarily translate to a high usefulness score for aiding decision making (see Table 7).

**Table 7.** Quality Assessment of Studies using CHEERS (16)

| **Study** | **Title/Abstract**<br>**(2 items)** | **Introduction**<br>**(2 items)** | **Methods**<br>**(10<sup>a</sup> items)** | **Results**<br>**(4<sup>b</sup> items)** | **Discussion**<br>**(1 item)** | **Disclosure**<br>**(2 items)** | **Total** | **% Items**<br>**met** |
| --- | --- | --- | --- | --- | --- | --- | --- | --- |
| *Cognitive* |  |  |  |  |  |  |  |  |
| Brown et al (18) | 1.5 | 2 | 6.5 | 2.5 | 1 | 2 | 15.5 | 74% |
| D'Amico et al (22) | 2 | 1.5 | 9 | 4 | 1 | 2 | 19.5 | 93% |
| Laakkonen et al (34) | 1.5 | 2 | 6.5 | 3.5 | 1 | 1 | 15.5 | 74% |
| Mervin et al (44) | 1.5 | 2 | 8 | 1 | 1 | 2 | 15.5 | 74% |
| Orgeta et al (23) | 1.5 | 2 | 8 | 3 | 1 | 2 | 17.5 | 83% |
| Sado et al (45) | 1 | 2 | 9 | 2 | 1 | 2 | 17 | 81% |
| Sogaard et al (42) | 2 | 2 | 8.5 | 2 | 1 | 2 | 17.5 | 83% |
| Spector et al (24) | 1.5 | 2 | 6 | 2 | 1 | 2 | 14.5 | 69% |
| Woods et al (25) | 1.5 | 1 | 8 | 4 | 1 | 2 | 17.5 | 83% |
| **Means** | 1.6 | 1.8 | 7.7 | 2.7 | 1.0 | 1.9 | 16.7 | 79% |
| *Training* |  |  |  |  |  |  |  |  |
| Ballard et al (26) | 1.5 | 2 | 4 | 3 | 1 | 2 | 13.5 | 64% |
| El Alili et al (37) | 2 | 2 | 8 | 3 | 1 | 2 | 18.0 | 86% |
| Livingston et al (27) | 1.5 | 2 | 8.5 | 3 | 1 | 1 | 17.0 | 81% |
| Meads et al (7) | 2 | 1.5 | 8.5 | 3 | 1 | 2 | 18.0 | 86% |
| Menn et al (30) | 2 | 2 | 8.5 | 4 | 1 | 1 | 18.5 | 88% |
| Romeo et al (21) | 1.5 | 2 | 9.5 | 1 | 1 | 1 | 16.0 | 76% |
| Van de Ven et al (38) | 1 | 2 | 7 | 1 | 0.5 | 2 | 13.5 | 64% |
| Williams et al (40) | 1 | 1.5 | 7 | 1.5 | 1 | 2 | 14.0 | 67% |
| **Means** | 1.6 | 1.9 | 7.6 | 2.4 | 0.9 | 1.6 | 16.1 | 76% |
| *Multi-disciplinary* |  |  |  |  |  |  |  |  |
| Henderson et al (48) | 1.5 | 2 | 9 | 3 | 1 | 2 | 18.5 | 88% |
| Jennings et al (41) | 1 | 1 | 6.5 | 3 | 0.5 | 1 | 13 | 62% |
| MacNeil Vroomen et al (39) | 1.5 | 2 | 9 | 4 | 1 | 1.5 | 19 | 90% |
| Michalowsky et al (32) | 2 | 2 | 8.5 | 1.5 | 1 | 2 | 17 | 81% |
| Mostardt et al (31) | 2 | 2 | 9.5 | 4 | 1 | 2 | 20.5 | 98% |
| Rädke et al (33) | 1.5 | 2 | 6 | 2 | 1 | 2 | 14.5 | 69% |
| Rosenvall et al (35) | 1 | 2 | 8.5 | 4 | 1 | 2 | 18.5 | 69% |
| Saxena et al (47) | 1 | 2 | 5.5 | 4 | 1 | 2 | 15.5 | 74% |
| **Means** | 1.4 | 1.9 | 7.8 | 3.2 | 0.9 | 1.8 | 17.1 | 79% |
| *Exercise* |  |  |  |  |  |  |  |  |
| D'Amico et al (20) | 2 | 2 | 10 | 3 | 1 | 2 | 20 | 95% |
| Davis et al (46) | 1.5 | 1.5 | 9 | 3 | 1 | 2 | 18 | 86% |
| Lamb et al (28) | 1.5 | 2 | 9 | 4 | 1 | 2 | 19.5 | 93% |
| Pitkala et al (36) | 1.5 | 2 | 8.5 | 2 | 1 | 2 | 17 | 81% |
| Sopina et al (43) | 1 | 1.5 | 7.5 | 3 | 1 | 0 | 14 | 67% |
| **Means** | 1.5 | 1.8 | 8.8 | 3.0 | 1.0 | 1.6 | 17.7 | 84% |
| *Other* |  |  |  |  |  |  |  |  |
| Tanajewski et al (19) | 1.5 | 2 | 9 | 4 | 1 | 2 | 19.5 | 93% |
| Howard et al (29) | 2 | 2 | 8.5 | 4 | 1 | 1 | 18.5 | 88% |
| **Means** | 1.8 | 2.0 | 8.8 | 4.0 | 1.0 | 1.5 | 19.0 | 90% |
| <sup>a</sup> 15 items assessed for Rosenvall et al modelling study |  |  |  |  |  |  |  |  |
| <sup>b</sup> 5 items assessed for Rosenvall et al modelling study |  |  |  |  |  |  |  |  |

### Cognitive Interventions

Nine studies explored interventions focused on cognition (Table 2). The majority used data from randomised controlled trials except one study with matched controls (45). All studies used treatment as usual (TAU) as the comparison group. Five studies used QALYs as the main measure of benefit (18,22,23,25,42) the remainder used patient health related quality of life (34), agitation (44), anxiety (24) and Criterion Time for Certification of Needed Long-Term Care (45). Study populations mainly comprised community dwelling PwD; studies also recruited nursing home residents (44,45) and a combination of community and nursing home residents (18).

The cognitive interventions in this review have been further categorised according to type: cognitive stimulation therapy, multi-component cognitive interventions and other cognitive therapies. Cognitive stimulation therapies were evaluated in three studies which all used a CUA approach and calculated QALYs (18,22,23). The studies comprised a CST programme for participants who had not received CST previously (23) and studies evaluating follow-up programmes of maintenance stimulation therapy (MST) (18,22). Follow-up periods ranged between six (18,22) and 25 months (23) (mean 12, SD 10.97). The first MST study reported an incremental cost-effectiveness ratio (ICER) of £26,835 per QALY, using proxy EQ5D assessments completed by caregivers (22). In this study cost-effectiveness acceptability curves (CEAC) demonstrated a 40% probability that MST would be seen as cost-effective at the £20,000 WTP threshold and 54% probability at £30,000. In the second MST study TAU dominated MST in terms of QALYs for a sub-group of patients living alone, with a probability of 55% cost-effectiveness at £20,000 WTP (18). Although this study rated highest in the review for quality assessed by CHEERS, it used a small secondary dataset which may not have been representative of the general population and was not powered to detect sub-group changes, therefore results should be interpreted with caution. The CST intervention did not improve cognition or quality of life for PwD, the TAU option performed better in terms of Alzheimer’s Disease Assessment Scale–Cognitive Subscale (ADAS-Cog) and was less costly so the TAU intervention dominated the CST intervention (23).

Multi-component cognitive interventions were also evaluated (42,45). The first study evaluated Learning Therapy, a combination of cognitive training and cognitive stimulation, in a nursing home population (45). The second assessed an early intervention approach which incorporated counselling, education and support in a community dwelling population (42). The Learning Therapy Trial conducted a CBA, matched controlled trial, whilst the second study with the early intervention approach used a CUA and a RCT design. Participants were followed up for significant periods of time at 12 months and 36 months respectively (42,45). Primary outcome measures differed, measuring time to institutionalisation (45) or QALYs (42).

The Learning Therapy study reported significant benefits at 12 months compared to TAU reporting $21 per month average cost-savings and an 88.8% probability that the intervention was cost-effective (45). However it should be noted that the matched control design was not gold-standard and such designs are at risk of introducing bias and confounding (49). The early intervention, multi-component, intervention was dominated by TAU in terms of QALYs (42).

The remaining four cognitive studies all evaluated different types of cognitive interventions. Woods et al used joint reminiscence therapy groups involving carers and PwD (25), Spector et al evaluated cognitive behavioural therapy (CBT) to reduce anxiety in PwD (24), Laakkonen et al used cognitive rehabilitation and involved promotion of self-management skills for people recently diagnosed with dementia (34), and Mervin et al assessed PwD interaction with a regular soft toy and an interactive toy with artificial intelligence (44). All studies used an RCT design, and CEA with the exception of Woods et al which used RCT design and CUA (25). With regard to primary outcomes Mervin et al measured agitation, Woods et al and Laakkonen et al evaluated quality of life and Spector et al used patient anxiety scores (24,25,34,44).

Two of these studies, by Laakkonen et al and Spector et al, proved to be cost-neutral (24,34). Laakkonen et al’s cognitive rehabilitation intervention study showed that participants in the intervention group showed significantly less decline in their verbal fluency scores compared with TAU, without increasing total costs (34). Similar improvements were seen for Clock Drawing Test scores. Spector et al’s CBT study reported significant improvements in depression which remained significant at six months; improvements in anxiety scores were also reported but failed to reach statistical significance (24). Although in the short-term the intervention reduced health and social care costs for the CBT group, overall, it was reported as cost-neutral.

The results of the Mervin et al study showed that there was little difference in ICER values/per Cohen-Mansfield Agitation Inventory - short form (CMAI-SF) improvement relative to TAU between the soft toy ($12.85) and interactive toy ($13.01) (44). No statistically significant between-group differences in agitation were reported at ten weeks and the study findings did not support the intervention being cost-effective. The Woods et al reminiscence intervention reported no significant difference in outcomes or service use between the intervention group and TAU, with an ICER = £2,586/per 0.597 unit change in Quality of Life in Alzheimer Disease (QoL-AD) score (25). Mean differences in QALYs between the two groups was negligible.

### Training Interventions

Eight studies evaluated cost-effectiveness of specialised dementia training for staff looking after PwD, to improve the quality of patient care and patient experience (Table 2). Seven of the eight interventions involved training for nursing home staff with one community-based training intervention for GPs. All studies were RCTs except for one cluster-randomized trial.

Williams et al evaluated a staff training communication intervention aimed at improving dementia behaviours in nursing homes using a cluster-randomized waitlist-controlled approach (40). El Alili et al evaluated a multi-component intervention for residents with advanced dementia (37). Livingston et al evaluated a staff training intervention to manage agitation (27). Romeo et al and Ballard et al evaluated interventions to reduce agitation by using a person-centred care intervention (21,26). Meads et al and Van de Ven et al both evaluated dementia care mapping (DCM) interventions in nursing homes (7,38) which involves observing an individual’s behaviour and then implementing an individualised care plan. A study to evaluate cost-effectiveness of GP training in dementia care and promote support groups and carer counselling was carried out by Menn et al (30).

With regard to study design El Alili et al, Livingston et al, Meads et al and Romeo et al all carried out CUAs, whilst Van de Ven et al used a CBA and Ballard et al, Menn et al and Williams et al undertook CEAs (7,21,26,27,30,37,38,40). The training intervention study with the longest follow up period was by Williams et al at 36 months (mean=15.4, SD 9.83) (40).

Four studies used QALYs (7,21,27,37) and one used quality of life assessed with a proxy Dementia Quality of Life (DEMQOL) measure (26). Van de Ven et al assessed health care consumption, falls, psychotropic drug use and staff absenteeism (38). Williams et al monitored percentage of time staff used ‘childish’ language with residents alongside the percentage of time residents were resistive to care (40). Menn measured time to institutionalisation (30). Meads et al and Van de Ven et al used similar methodology; both used patient assessment and observation to evaluate their DCM studies, using agitation as the primary outcome and a healthcare provider/services perspective (30,38).

Several training interventions showed improvements in primary outcomes. The Williams et al staff communication intervention reduced resistiveness to care behaviours and reported an ICER of US$4.31 per resident per one percentage point reduction in resistiveness to care (40). This study was limited by lack of appropriate statistical analysis, no ICER in terms of QALYs was reported making direct comparison with other studies difficult. A strength of this study was its long time-horizon of 36 months; however it is limited by the small sample size and generalisability is limited due to the small geographical location of the nursing homes. Ballard et al showed a statistically significant improvement of 2.54 points in quality of life (mean difference, DEMQOL proxy) for the patient centred care intervention compared to TAU (26). Statistically significant benefits were also demonstrated for Cohen-Mansfield Agitation Inventory (CMAI) and Neuropsychiatric Inventory Nursing Home Version outcomes compared to TAU; cost savings were reported for this multi-component intervention. The perspective of the study and it’s relation to the costs being evaluated was unstated which was a limitation. Romeo et al’s person-centred care based intervention also demonstrated clinically significant benefits with reduction in agitation and improvement in quality of life, when used with people with clinically significant agitation living in nursing homes (21). The Ballard et al intervention was dominant compared to TAU in terms of QALYs gained; when used in combination with TAU it demonstrated 100% probability of cost-effectiveness at a WTP threshold of £70,000 compared with TAU alone (26).

The multi-component intervention evaluated by El Alili et al was reported to dominate TAU in terms of QALYs; a strength of this intervention was its one-year time horizon (37). The Van de Ven et al DCM intervention was evaluated as cost-neutral, and the number of outpatient hospital appointments was reduced compared to usual care (38). Analysis of uncertainty was not stated and no ICER was presented.

The Livingston et al staff training intervention was cost-effective in terms of QALYs and reported an ICER of £14,064 per QALY gained with a 62% probability of cost-effectiveness at a WTP of £20,000 per QALYs (27). The authors report that it was not effective in terms of improving the primary outcome of agitation measured by CMAI scores between the intervention group and TAU.

Despite having the longest of all follow up periods at 48 months, the Menn et al GP training intervention showed no significant reduction in time to institutionalisation (30). The Meads et al intervention was not cost-effective in terms of QALYs, ICER=£64,380 per QALY gained (7). Small improvements were achieved in reducing agitation measured by CMAI, but it failed to reach statistical significance.

### Multi-disciplinary Interventions

Eight articles evaluated multi-disciplinary dementia care programmes. Interventions have been further categorised according to type: dementia care management, case management, and other multi-disciplinary dementia care (meeting centres and a primary care dementia clinic were separately evaluated in two studies).

Dementia care management evaluations by Jennings et al, Michalowsky et al and Rädke et al used multi-disciplinary care in the community to assess and support patients (32,33,41). Rosenvall et al evaluated three interventions: care management, family support and multi-disciplinary rehabilitation; modelling potential cost savings of each (35). Study designs varied, with RCT (32,41) a cluster RCT (33) and a modelling approach being taken (35). Rädke et al undertook a CEA and Michalowsky et al performed a CEA using the same participant dataset (32,33). Rädke et al undertook sub-group analysis to determine which group of patients benefitted most from this type of programme (33). Jennings et al had the largest sample size (n=3269) not just in this category but across all intervention categories (41).

The dementia care management interventions evaluated by Michalowsky et al and Rädke et al, dominated TAU in terms of QALYs (32,33). Both used QALYs derived from the 12-Item Short Form Survey (SF-12) quality of life measure as their primary outcome. Compared to TAU the Michalowsky et al dementia care management intervention was reported to increase QALYs (by an average of 0.05), reduce hospitalisation and delay the transfer to long-term care (seven months) and costs were decreased (by an average of €569) (32). Cost-effectiveness probability was 88% at WTP thresholds of €40,000/QALY gained and was higher in patients who lived alone compared to those not living alone (96% versus 26%).

Rädke et al reported a probability of cost-effectiveness at 88% at a WTP threshold of €40,000 per QALY (33). For the sub study populations, higher probability of cost-effectiveness was reported in older PwD (<80 years) than younger (>80 years) at the same WTP threshold (87% versus 48% respectively). Probability of cost-effectiveness was very high in females compared to males (96% versus 16%), in PwD living alone compared to those not living alone (96% versus 26%) and in patients with more co-morbidity than less comorbidity (96% versus 26%). The authors suggest that the high probability of cost-effectiveness in females and those living alone could be attributed to these groups having fewer relatives or carers to provide care and support and therefore having a higher number of unmet needs, meaning that they are more likely to benefit from a multi-disciplinary management programme. The intervention was more effective in those with a higher functional deficit and in patients with moderate to severe cognitive impairment than those without cognitive impairment.

Jennings et al reported a hazard ratio of 0.60, demonstrating that dementia care management programme participants were less likely to be admitted to a long-term care facility than with TAU (41). The authors concluded that the intervention was cost neutral.

Rosenvall et al’s modelling simulation concluded that cost savings were possible with care management, family support and multidisciplinary rehabilitation as they delayed admission time to long-term care (35). The estimated time taken to reach economic break-even points was based on the estimate of how long a patient’s transition to long-term care could potentially be delayed by. For care management this was 2.8 days, for structured family support 1.8 days, for rehabilitative cognitive, physical and social activation the break-even point was 43.0 days. Rehabilitation was reported to be most cost-effective in the severe phases of Alzheimer’s disease. Potential cost savings were demonstrated through saving costs on long term care by delaying the decline in cognition and social functioning of PwD and subsequently the time of transition to long term care.

Case management, supporting the person with dementia and their carer in conjunction with a multidisciplinary team, was evaluated by MacNeil Vroomen et al and Mostardt et al (31,39). MacNeil Vroomen et al undertook a CUA with an observational design and a societal perspective, whilst Mostardt et al evaluated CEA in a non-randomised matched controlled trial from a healthcare provider perspective (31,39). Mostardt et al reported an ICER of €53/per additional month in a home environment (31). The difference in average additional months spent in the home environment was significant at 16.14 months for the intervention group patients compared with 12.2 months for the control group patients (p = 0.02). However, it was noted that no limitations were discussed in this study and they used a non-randomised matched controlled study design. MacNeil Vroomen et al reported that the intensive care management model (ICMM) intervention was cost-effective compared to the linkage model and TAU (39). The difference in QALYs gained between ICMM and control was non-significant. The probability that ICMM was cost-effective compared to control was 99% at WTP of €30,000/QALY and dominated TAU in terms of QALYs. However, no sensitivity analysis was conducted to test the robustness of the outcome. Additionally, the observational, non-randomised design may have introduced selection bias and more heterogeneity into the study population.

Henderson et al conducted a non-randomised study to evaluate meeting centres providing day support using CUA (48). The study had a short time horizon at six months and was not cost-effective in terms of QALYs. When using QOL-AD as the primary outcome, the probability of cost-effectiveness was 50% at a WTP of €5,000 for a one-point increase.

Saxena et al also undertook an evaluation of a multi-disciplinary intervention comparing a primary care dementia clinic (PCDC) with a hospital-based memory clinic (MC) using a non-randomised quasi-experimental design and CUA (47). QALYs were higher for the PCDC group and the ICER at 12 months was S$29,042/per QALY which was less than the assumed threshold of S$78,690. The authors concluded that the care provided by the PCDC had similar effectiveness to that provided by a hospital memory clinic and it was more effective than that provided by the other polyclinics evaluated so the authors suggest that these clinics could be cost-effectively set up elsewhere in primary care.

### Exercise Interventions

Five articles looked at the effect of physical activity on people with dementia. Although all studies were randomised controlled trials, they employed various economic evaluation methods. Lamb et al, Sopina et al and D’Amico et al carried out a CUA (20,28,43), and Davis et al and Pitkala et al used a CEA (36,46). Only one intervention included people living in nursing homes (20), the rest focused on people living in the community. The majority of the studies focused on group-based exercise outside of the home.

None of the five exercise studies evaluated demonstrated cost-effectiveness in terms of QALYs, however, improvements in primary outcomes were seen in two of the studies (36,46). Davis et al evaluated group resistance training classes and aerobic training classes and compared these to group balance and tone classes which had not been specifically designed to target cognitive decline (46). They reported that the resistance and aerobic training classes dominated the balance and tone classes at six months in terms of seconds gained/lost on Stroop test of cognitive function and were less costly. In Pitkala et al’s study, both the home exercise and group exercise intervention groups demonstrated significantly slower decline in functioning measured by Functional Independence Measure (FIM) (−7.1, −10.3 FIM change respectively) than the control group (−14.4 FIM change) without increasing the total costs of health and social services (36).

### Other Interventions

Tanajewski et al undertook a CUA with RCT design looking at developing a specialist unit to provide care for confused patients admitted to a general hospital (19). During a follow up period of three months the intervention was found to dominate TAU in terms of QALYs with a probability of cost-effectiveness of 81% at a WTP threshold of £20,000/QALY.

Howard et al carried out an RCT employing a CUA to evaluate telecare and assistive technology from both a health and social care and a societal perspective (29). The intervention was not cost-effective in terms of QALYs from either a health and social care or societal perspective and it did not enable people with dementia to live safely at home for longer.

#### Review of Reviews

Alves et al undertook a systematic review of five RCT’s on the efficacy and feasibility of cognitive interventions for those with Alzheimer’s Disease (9). The review only included cognitive stimulation, cognitive training and cognitive rehabilitation. The review was rated as critically low for confidence in quality using AMSTAR 2 with no evidence of investigation of publication bias and its potential impact on results. Only one relevant economic study of cost-effectiveness was identified: a programme of cognitive stimulation for PwD living either at home or in nursing homes, not receiving cholinesterase inhibitor treatment (participant numbers not stated). The intervention was not reported as cost-effective.

A narrative review was undertaken by Banerjee into the efficacy of providing memory services (50), due partly to the non-systematic approach the review was rated as critically low for confidence in quality using AMSTAR 2. The author referred to a simulation model undertaken by Banerjee and Wittenberg which modelled the effectiveness of implementing a national memory service provision to prevent care home admissions (51). They estimated cost savings to society which increased over the ten-year period as the number of those admitted to care homes was reduced. Cost-effectiveness was reported at £20,000 per 0.02 QALY.

Home support interventions for PwD living in the community were systematically reviewed by Clarkson et al (13). The review included 14 economic evaluations, six of these evaluated only carer outcomes and as such were excluded here. The Pitkala et al study was reviewed, it has already been reviewed in this paper and will be not be reviewed again here to avoid duplication (36). The remaining seven relevant economic evaluations looked at the following interventions: care management (n=2), group living, occupational therapy, activity sessions, institution-based care and specialist dementia day care compared to usual care at home. Four of the evaluations used CEA, two used CUA and one used cost-consequence analysis. Only occupational therapy was reported to show cost-effectiveness. Confidence in the Clarkson et al review was rated as critically low for confidence in quality using AMSTAR 2, risk of bias was not discussed in the review results.

As part of a systematic review of interventions to reduce agitation in people with dementia in any setting, Livingston et al identified two economic studies (11). Mintzer et al evaluated a programme to reduce hospitalisation using a blended inpatient/outpatient intervention and Norman et al evaluated a comparison of DCM and person-centred care (52,53). Both studies measured economic outcomes using CMAI scores. Mintzer et al recorded a 0.89 change in total CMAI for $1000 expenditure for the inpatient/outpatient intervention versus only 0.27 for the inpatient programme (52). This suggests effectiveness but as cost-effectiveness thresholds are not known it is not possible to determine cost-effectiveness. Norman et al reported that for person-centred care relative to usual care there were costs of A$6.43 per CMAI point averted, for the DCM intervention the costs were higher at A$46.89 (53). The summary economic measure for this study was cost per CMAI score change, further data on effectiveness of this intervention was not available within the review.

Additionally, as part of the review Livingston et al created a simulation model using the most effective strategies identified from an effectiveness review of 30 studies of wide-ranging dementia therapies and existing patient cohort data (11). Modelling of a multi-component intervention for participants with mild to moderate dementia revealed 82% probability of cost-effectiveness at a maximum willingness to pay threshold of £20,000/QALY. However due to the multi-component nature it was not possible to determine which particular component of the intervention was most effective. The review rated moderate for confidence in quality using AMSTAR 2.

The Nickel et al review (3) contained three studies on physical activity (20,36,46) and six on cognitive interventions (22–25,34,42,54) which were relevant to this systematic review. The studies contained in this review have already been individually identified through the literature review, and methods and results are discussed above. The review rated low for confidence in quality using AMSTAR 2 as assessment of risk of bias was not detailed.

## Discussion

### Summary of main findings

This paper reviewed economic evaluations of both hospital-based and community interventions for dementia and MCI. A wide range of interventions from researchers around the world was evaluated for patients both at home and in nursing homes.

Of the 37 studies and five reviews evaluated in this paper 13 demonstrated evidence to favour interventions, although some of these had limitations. The intervention category with the strongest evidence of cost-effectiveness was multi-disciplinary interventions. Four of the eight multi-disciplinary interventions were likely to be cost-effective in terms of QALYs gained compared to TAU. Dementia care management was the multi-disciplinary intervention which showed strongest evidence of cost-effectiveness, two of the four studies demonstrated cost-effectiveness in terms of QALYs (32,33) and two reported delays in admission to long-term care (35,41).

In the cognitive intervention category, although short term CST was not shown to be cost-effective, MST demonstrated evidence of cost-effectiveness, although only two economic studies were available. As previously mentioned, group CST is already recommended as an intervention (8) and has been proven to be cost-effective in a large-scale study outside of the timeframe of this review (55). Both MST studies evaluated as part of this review were cost-effective in terms of QALYs compared to TAU. Overall, the cognitive interventions did not demonstrate evidence of cost-effectiveness. The economic evidence was limited due to low numbers of studies per intervention making it impossible to make an informed decision about their cost-effectiveness. In Clarkson et al’s review occupational therapy was evaluated as cost-effective but this was the only study which reviewed this intervention.

There was also evidence for care home and nursing home staff training interventions. Three out of eight studies favoured the intervention over TAU and found cost-effectiveness in terms of QALYs or showed significant patient benefits. A fourth study was cost neutral.

Limitations were identified in some of the studies and evidence was weakened by small number of studies per intervention, small sample sizes, short timeline or reliability of evidence. Four of the studies reporting cost-effectiveness did not use an RCT design and this could have led to biased estimates of effect (31,39,45,47). The generalisability of the sample to the wider population was a potential issue in two studies as the study population was limited to a rural community (32,33). Three of the six multi-disciplinary studies took place in the same country, which may lead to a geographical bias (31–33). It should also be noted that only one of the multi-disciplinary studies assessed UK based services (48).

### Limitations

Despite the broad search terms used it is possible that some economic evaluation studies may have been missed. There may be an element of publication bias as it is acknowledged that authors may be less willing to submit studies for publication that don’t demonstrate cost-effectiveness or cost-savings.

Synthesising evidence from studies and reviews evaluating a wide range of interventions presented challenges. Due to the variety of outcome measures used, the heterogeneity of the study methods and the variety of different interventions it was difficult to compare the cost-effectiveness of different interventions. The lack of WTP thresholds for different countries made it difficult to compare the ICERs between countries.

The review was also hampered by lack of robust economic evidence generally for non-pharmacological studies. This in turn resulted in limited evidence for each category of intervention.

The narrative review by Banerjee was assessed as critically low using AMSTAR 2, however, it is acknowledged that the AMSTAR 2 tool was designed for use with systematic reviews and that this may account for the low quality score. However, non-systematic reviews can still be valuable and informative and can generate novel insights by allowing for a flexible and adaptable approach to article selection (56).

### Recommendations for Future Research

The results of the review demonstrates gaps in the economic evidence on non-pharmacological interventions which could benefit from further research. There was a lack of economic studies on the cost-effectiveness of creative therapies such as art, music, drama, creative writing and dance and also sensory therapies such as aromatherapy and massage. There was limited evidence shown for multi-component cognitive studies and CBT. Further high-quality research would evaluate whether these interventions are cost-effective.

## Conclusion

The intervention with the strongest evidence cost-effectiveness was dementia care management. There was also evidence to suggest cost-effectiveness of MST and occupational therapy, although evidence was limited by availability of studies. More economic evidence on the cost-effectiveness of dementia care interventions is needed, with consistency around measurement of costs and outcomes data to inform policy and decision makers future decision making. Better information and higher-quality studies are also needed in order to improve decision makers’ confidence to promote cost-effective dementia interventions in the future.

## Supporting information

Prisma S

## Data Availability

All data produced in the present work are contained in the manuscript

## Contributions and Support

GE, PM and RP contributed to the conception and design of the study and methodology; GE and PM contributed to the development of the protocol. Searches and screening were carried out by GE, PM and EG. Data extraction, and quality appraisal were conducted by GE and EG. GE drafted the manuscript. PM, RP, CS and EG critically reviewed the manuscript and revised it critically for important intellectual content; all authors approved the final version to be published.

## Support acknowledgment

This research was supported by the National Institute for Health Research (NIHR) Applied Research Collaboration Kent, Surrey, Sussex. The views expressed are those of the author(s) and not necessarily those of the NHS, the NIHR or the Department of Health and Social Care.

## Data Availability

Data availability is not applicable to this article as no new data were created or analysed in this review.

