## Supplementary material for "Effectiveness of community non-pharmacological interventions for mild cognitive impairment and dementia: a systematic review of economic evaluations and a review of reviews": Prisma S

PRISMA-S Checklist

| Section/topic | # | Checklist item | Location(s) reported |
| --- | --- | --- | --- |
| Information sources and methods | | | |
| Database name | 1 | Name each individual database searched, stating the platform for each. | Search strategy p4 |
| Multi-database searching | 2 | If databases were searched simultaneously on a single platform, state the name of the platform, listing all of the databases searched | N/A |
| Study registries | 3 | List any study registries searched | N/A |
| Online resources and browsing | 4 | Describe any online or print source purposefully searched or browsed (e.g. tables of contents, print conference proceedings, web sites) and how this was done | N/A |
| Citation searching | 5 | Indicate whether cited references or citing references were examined and describe any methods used for locating cited/citing references (e;g. browsing reference lists, using a citation index, setting up email alerts for references citing included studies). | Search strategy p4 |
| Contacts | 6 | Indicate whether additional studies or data were sought by contacting authors, experts, manufacturers, or others | N/A |
| Other methods | 7 | Describe any additional information sources or search methods used | N/A |
| Search strategies | | | |
| Full search strategies | 8 | Include the search strategies for each database and information source, copied and pasted exactly as run | Search strategy p4 |
| Limits and restrictions | 9 | Specify that no limits were used, or describe any limits or restrictions applied to a search (e.g. date or time period, language, study design) and provide justification for their use. | Search strategy p4 |
| Search filters | 10 | Indicate whether published search filters were used (as originally designed or modified) and if so, cite the filters used. | N/A |
| Prior work | 11 | Indicate whether search strategies from other literature reviews were adapted or re used for substantive part or all of the search, citing the previous review (s) | N/A |
| Updates | 12 | Report the methods used to update the search(es) (e.g. rerunning searches, email alerts) | N/A |
| Dates of searches | 13 | For each search strategy, provide the date when the last search occurred | Search strategy p4 |
| Peer review | | | |
| Peer review | 14 | Describe and search peer review process | Study selection p4 |
| Managing records | | | |
| Total records | 15 | Document the total number of records identified from each database and other information sources | Figure 1 page 19 |
| Deduplication | 16 | Describe the processes and any software used to deduplicate records from multiple database searches and other information sources | Results p5 |
